# Assessing regional intracortical myelination in schizophrenia spectrum and bipolar disorders using the optimized T1w/T2w-ratio

**DOI:** 10.1101/2023.06.02.23290882

**Authors:** Kjetil Nordbø Jørgensen, Stener Nerland, Nora Berz Slapø, Linn B. Norbom, Lynn Mørch-Johnsen, Laura Anne Wortinger, Claudia Barth, Dimitrios Andreou, Ivan I. Maximov, Oliver M. Geier, Ole A. Andreassen, Erik G. Jönsson, Ingrid Agartz

## Abstract

**Background:** Dysmyelination could be part of the pathophysiology of schizophrenia spectrum (SCZ) and bipolar disorders (BPD), yet few studies have examined myelination of the cerebral cortex. The ratio of T1- and T2-weighted magnetic resonance images (MRI) correlates with intracortical myelin. We investigated the T1w/T2w-ratio and its age trajectories in patients and healthy controls (CTR) and explored associations with antipsychotic medication use and psychotic symptoms.

**Methods:** Patients with SCZ (n=64; mean age = 30.4 years, SD=9.8), BPD (n=91; mean age 31.0 years, SD=10.2), and CTR (n=155; mean age = 31.9 years, SD=9.1) who participated in the TOP study (NORMENT, University of Oslo, Norway) were clinically assessed and scanned using a General Electric 3T MRI system. T1w/T2w-ratio images were computed using an optimized pipeline with intensity normalization and field inhomogeneity correction. Vertex-wise regression models were used to compare groups and examine group × age interactions. In regions showing significant differences, we explored associations with antipsychotic medication use and psychotic symptoms.

**Results:** No main effect of diagnosis was found. However, age slopes of the T1w/T2w-ratio differed significantly between SCZ and CTR, predominantly in frontal and temporal lobe regions: Lower T1w/T2w-ratio values with higher age were found in CTR, but not in SCZ. Follow-up analyses revealed a more positive age slope in patients who were using antipsychotics and patients using higher chlorpromazine-equivalent doses.

**Conclusions:** While we found no evidence of reduced intracortical myelin in SCZ or BPD relative to CTR, different regional age trajectories in SCZ may suggest a promyelinating effect of antipsychotic medication.

## 1. Introduction

Schizophrenia spectrum and bipolar disorders are severe mental disorders that affect more than 1% of the population (Perälä et al., 2007). These disorders are proposed to exist along a psychosis continuum (Pearlson, 2015; Tamminga et al., 2014). Psychotic symptoms are also common in bipolar disorders, with an estimated lifetime prevalence above 60% in bipolar I disorder and above 20% in bipolar II disorder (Aminoff et al., 2022; van Bergen et al., 2019). Current evidence favors the view that both disorders reflect dysconnectivity within and across several brain circuitries rather than having focal origin (Friston et al., 2016; Friston and Frith, 1995; Kelly et al., 2018; Xia et al., 2019). Dysmyelination has been proposed as one possible mechanism (Bartzokis, 2002; Whitford et al., 2012). This hypothesis is supported by genetic (Goudriaan et al., 2014; Hakak et al., 2001) and post-mortem studies (Kolomeets and Uranova, 2018; Uranova et al., 2011; Vikhreva et al., 2016) indicating lower myelin content as well as lower density and altered morphology of myelinating oligodendrocytes in the prefrontal cortex (Kolomeets and Uranova, 2018; Uranova et al., 2011; Vikhreva et al., 2016). Notably, the maturation of myelin in association fiber tracts and frontal regions of the cerebral cortex, regions known to be involved in psychotic disorders, extends into early adulthood, which is a period of heightened incidence of psychosis (Paus et al., 2008; Whitford et al., 2012).

Magnetic resonance imaging (MRI) studies on dysmyelination in psychotic disorders have mainly examined white matter, whereas few have examined intracortical myelination (Ganzetti et al., 2015; Iwatani et al., 2015; Wei et al., 2022; Wei et al., 2020). In a previous study, we examined the cortical gray-white matter contrast (GWC), i.e., the contrast between T1-weighted (T1w) intensities in gray matter and adjacent superficial white matter, which is inversely correlated with intracortical myelin. We found higher GWC values in sensory and motor regions in patients with schizophrenia spectrum disorders and, to a lesser extent, in bipolar disorders compared to healthy controls (Jorgensen et al., 2016). More recently, Makowski et al (2019) used structural covariance and principal component analysis on the GWC in patients with first-episode psychosis. They reported a similar trend-level difference in a GWC component representing sensory and motor regions in patients relative to healthy controls. However, the GWC is an indirect measure, and it is necessary to validate these findings using other measures of intracortical myelin.

In bipolar I disorder, Sehmbi et al (2018) found positive associations between a T1w intensity-based measure of intracortical myelin and verbal memory. They also reported an inverted U-shaped relationship with age in healthy controls but not in patients with bipolar I disorder (Sehmbi et al., 2019). Higher T1 relaxation times in bipolar disorders have been found using quantitative MRI (Rangel-Guerra et al., 1983) and in one study this was found in regions including the somatosensory and temporal cortices (Necus et al., 2019).

Myelin may also be involved in the therapeutic action of antipsychotic medications (Bartzokis, 2012; Kroken et al., 2014). In animal studies, a lipogenic effect of antipsychotic agents has been demonstrated (Ersland et al., 2017; Ferno et al., 2011), and in clinical studies an association between serum lipid levels and treatment response was reported (Gjerde et al., 2018a; Kim et al., 2019; Procyshyn et al., 2007). Notably, Tishler et al (2018) investigated a measure of frontal lobe intracortical myelin volume and found a significant association within the first year of antipsychotic medication exposure that declined with prolonged exposure. In a previous study, users of second-generation antipsychotics had a higher intracortical myelin volume compared with users of first-generation antipsychotics (Bartzokis et al., 2007). While these findings are based on an indirect measure, they provide *in vivo* evidence suggestive of a promyelinating effect of antipsychotic medication in the frontal lobe of the cerebral cortex.

The T1w/T2w-ratio has been proposed as a measure of intracortical myelin (Glasser et al., 2016; Glasser and Van Essen, 2011). This interpretation is based on observations that the T1- and T2-weighted (T2w) MRI signals have positive and negative correlations with myelin content, respectively, such that the contrast due to myelin is enhanced in the ratio (Koenig, 1991; Koenig et al., 1990). Furthermore, shared field inhomogeneities in the T1w and T2w images are attenuated (Glasser and Van Essen, 2011). The T1w/T2w-ratio has been studied in neurological disorders such as multiple sclerosis (Beer et al., 2016), Huntington’s disease (Rowley et al., 2018), and Alzheimer’s disease (Pelkmans et al., 2019). In patients with schizophrenia spectrum disorders, Iwatani et al (2015) reported lower global T1w/T2w-ratio means both in gray and white matter compared to healthy controls, but no voxel-wise group differences in the cerebral cortex. Ganzetti et al (2015) used non-brain intensities to calibrate the T1w/T2w-ratio and found lower regional gray matter values in patients with schizophrenia spectrum disorders, particularly in the temporal lobe, frontal lobe, and the insula. However, two recent studies have indicated a more complex layer-dependent pattern of changes in first-episode psychosis (Wei et al., 2022; Wei et al., 2020). In bipolar disorders, Ishida et al (2017) reported lower T1w/T2w-ratio values in white matter regions relative to healthy controls, but no significant differences in gray matter.

Between-subject comparisons of the T1w/T2w-ratio are challenging due to the non-quantitative nature of MRI signal intensities. In a previous study, we evaluated the measurement properties of 33 T1w/T2w-ratio processing pipelines (Nerland et al., 2021). Correction for field inhomogeneities improved the agreement with the expected myeloarchitecture (i.e., the expected distribution of myelin across cortical areas). Furthermore, intensity normalization ensured acceptable test-retest reliability, which is of particular importance for between-subject comparisons.

In the present study, we investigated cortical T1w/T2w-ratio values in patients with schizophrenia spectrum and bipolar disorders relative to healthy controls. An optimized intensity normalized pipeline was used for computing the T1w/T2w-ratio maps with corrections for partial volume effects, surface outliers, and field inhomogeneities (Nerland et al., 2021). We examined if T1w/T2w-ratio values or age trajectories differed between each patient group and healthy controls with the following two hypotheses: First, that T1w/T2w-ratio values would be lower in primary sensory and motor regions in both patient groups. Second, that use of antipsychotic medication would, particularly in frontal regions of the cerebral cortex, be positively associated with the T1w/T2w-ratio. In exploratory analyses, we examined associations with psychotic symptoms.

## 2. Methods

### 2.1 Study design

Participants were recruited from hospitals in the greater Oslo region to the Thematically Organized Psychosis (TOP) study conducted by the Norwegian Centre for Mental Disorders Research (NORMENT). Patients who met the criteria for a DSM-IV schizophrenia spectrum disorder (SCZ), including schizophrenia, schizophreniform disorder and schizo-affective disorder, or bipolar disorders (BPD), including bipolar I disorder, bipolar II disorder and bipolar disorder not otherwise specified, were included in the current study. Healthy controls were randomly drawn from the national population registry in the same geographical region and asked to participate. The study complied with the Helsinki Declaration and was approved by the Regional Committee for Medical Research Ethics (REC South-East Norway) and the Norwegian Data Inspectorate. All participants gave informed consent.

The inclusion criteria for the TOP study include age between 18-65 years, no mental disability (defined as IQ<70), no history of head trauma with loss of consciousness, and no neurological disorder or other organic disorder thought to affect brain function.

Healthy controls were screened using the PRIME-MD (Spitzer et al., 1994). The absence of a mental disorder, substance use disorder, and history of severe mental disorders among first-degree relatives were criteria for inclusion. Healthy controls were selected from a larger pool (n=278) based on age- and sex-matching to the patient sample (SCZ and BPD). Matching was performed with the MatchIt package in R (version 4.2.3; R Core Team, 2018) (Ho et al., 2011). One-to-one matching was performed with the nearest neighbor method and quantile-quantile (Q-Q) plots were inspected to ensure adequate matching.

### 2.2 Diagnostic and clinical assessment

Clinical assessments were conducted by trained physicians, psychiatrists, or clinical psychologists. Diagnoses were verified using the Structured Clinical Interview for the DSM-IV Axis I disorders (SCID-IV) (Spitzer et al., 1992). Current symptoms were rated using the Positive and Negative Syndrome Scales (PANSS) (Kay et al., 1987). Scores for positive, negative, disorganized, excited, and depressed symptoms were calculated according to the five-factor model by Wallwork et al (2012). Current medication use was obtained by interview or chart review and included information on antipsychotic, antiepileptic, antidepressant, and anxiolytics/hypnotic medication. For each medication, type and dose were recorded. Antipsychotic medication dosages were converted to chlorpromazine equivalent doses (CPZ) (Andreasen et al., 2010). Intelligence quotient (IQ) was assessed with the Wechsler Abbreviated Scale of Intelligence (WASI-II) (Wechsler, 2007), and general psychosocial functioning was rated using the split version of the Global Assessment of Functioning Scale (GAF) (Pedersen et al., 2007). The median time from clinical assessment (defined as the day of the PANSS interview) to MRI acquisition among patients was 12 days, with an interquartile range of 7 – 25 days.

### 2.3 MRI acquisition

Patients and healthy controls were scanned using a 3T General Electric Discovery MR 750 system, equipped with a 32-channel head coil, between 2015 and 2019. T1w and T2w sequences were both acquired with 1 mm isotropic resolution. The T1w sequence was a 3D inversion recovery-prepared fast spoiled gradient echo recall (BRAVO) sequence with the following parameters: Repetition time (TR) = 8.16 ms; Echo time (TE) = 3.18 ms; Inversion time (TI) = 450 ms; Flip angle = 12°; Bandwidth = 244 Hz/px; ARC = 2; Acquisition time (TA) = 04:43. The T2w sequence was a 3D fast spin echo (CUBE) sequence with the following parameters: TR = 2500 ms; TE = 71.68 ms; FA = 90°; Bandwidth = 488 Hz/px; Echo train length (ETL) = 100; ARC = 2x2; TA = 04:23. Phased array uniformity enhancement (PURE) was enabled for both sequences. MRI images were inspected by a neuroradiologist and excluded if pathological findings were present.

### 2.4 MRI post-processing

FreeSurfer (v6.0.0; https://surfer.nmr.mgh.harvard.edu/) was used to reconstruct cortical surfaces, representing the boundary between gray and white matter (i.e., the inner gray-white surface of the cortex) and between gray matter and cerebrospinal fluid (i.e., the outer surface of the cortex or ‘pial surface’), based on T1w images.

FreeSurfer is open source and has been described in detail previously (Fischl, 2012). Reconstructed surfaces were visually inspected and edited according to standard guidelines. Images were excluded in the event of substantial motion artifacts or otherwise poor image quality.

### 2.5 Calculation of the T1w/T2w-ratio

To compute the T1w/T2w-ratio, we rigidly registered T2w images to T1w images using *bbregister* in FreeSurfer with FSL initialization (Greve and Fischl, 2009). We then applied N4ITK field bias correction and normalized intensities with the WhiteStripe algorithm. The T1w image was then divided by the T2w image to form the T1w/T2w-ratio, which was corrected for partial volume effects (Shafee et al., 2015). Next, T1w/T2w-ratio voxel values were projected onto the gray-white surface by sampling along layers representing equivolumetric distances of 10% to 80% of the vertex-wise cortical thickness (Waehnert et al., 2016). Finally, we performed surface-based outlier correction based on a previously published approach (Glasser and Van Essen, 2011). This pipeline was shown in a previous study to be robust to the presence of field inhomogeneities and to improve test-retest reliability whilst preserving inter-individual variation (Nerland et al., 2021).

We visually inspected each T1w/T2w-ratio map. If the maps deviated from known myeloarchitecture, the T1w and T2w volumes were inspected. If artefacts or low image quality were found in either of the scans, the participant was excluded.

### 2.6 Statistical analyses

Group differences in demographical and clinical variables were assessed using analysis of variance (ANOVA) or *Χ*^2^ tests with *post hoc* Bonferroni tests where appropriate.

In the primary analyses, we fitted age- and sex-adjusted vertex-wise (i.e., one model per vertex on the fsaverage gray-white surface) general linear models (GLMs) to examine 1) main effects of diagnosis (SCZ versus CTR, BPD versus CTR) and 2) diagnosis × age interaction effects. T1w/T2w-ratio maps were concatenated and smoothed (10 mm FWHM) before running the analyses. The models were then specified using the FreeSurfer command *mris_glmfit* with two categorical factors, diagnosis (three levels) and sex (two levels), and age as a continuous variable. Cluster-wise correction for multiple testing was employed with a cluster-forming threshold of 0.001 and cluster-wise probability of 5%. Correction for analysis across two hemispheres was applied. If significant diagnosis × age interaction effects were found, we further examined if the age slope differed from zero within each diagnostic group separately. The latter analyses were regarded as follow-up examinations, and we chose a liberal p-value threshold of p<0.01.

Further, we aimed to examine whether T1w/T2w-ratio values were associated with antipsychotic medication use or psychotic symptoms in regions where significant differences were found in the primary analyses. We first extracted the mean T1w/T2w-ratio values for each significant cluster. These were defined as dependent variables. To examine associations with antipsychotic medication use, antipsychotic medication status (current use/no use) as well as medication status × age interaction terms were entered as predictors of interest in the first set of models conducted in the patient sample (n=155). In the second set of models, we examined only patients who were using antipsychotic medication (n=86) and entered CPZ and CPZ × age interaction terms as predictors. We then performed separate analyses among all patients (n=155) using the PANSS total score and each of the five Wallwork factor scores as predictors of interest. All models were adjusted for age, sex, and diagnosis (SCZ/BPD). The analyses were conducted in SPSS version 28.

## 3. Results

### 3.1 Description of the study sample

The study sample consisted of 64 patients with SCZ, 91 patients with BPD and 155 CTR. The mean age did not differ between the groups. The sex distribution differed between groups, and *post hoc* tests indicated a non-significant trend towards more males in the SCZ group and more females in the BPD group.

The distributions of other demographic and clinical variables are shown in Table 1. Briefly, years of education and estimated IQ differed between the groups. Patients with SCZ had higher current PANSS positive, negative, and disorganized symptom scores, but not excited or depressive symptom scores compared to patients with BPD. Patients with SCZ also had lower GAF scores compared to patients with BPD.

**Table 1.** Sample characteristics.

| Sample characteristics | SCZ (n=64) | BPD (n=91) | CTR (n=155) | Statistics | Post hoc tests |
| --- | --- | --- | --- | --- | --- |
| *Demographic* |  |  |  |  |  |
| Age, years (M, SD) | 30.4 (9.8) | 31.0 (10.2) | 31.9 (9.1) | $F = 0.6$, n.s. | |
| Sex (male, %) | 37 (58%) | 34 (37%) | 73 (47) | $\chi^2 = 6.4$, $p=0.04$ | |
| Education, years (M, SD) | 12.6 (2.2) | 13.6 (2.1) | 14.8 (2.0) | $F = 27.1$, $p<0.001$ | SCZ < BPD, CTR. BPD < CTR. |
| WASI IQ (M, SD) | 104.9 (13.3) | 111.9 (11.6) | 115.4 (11.9) | $F = 19.0$, $p<0.001$ | SCZ < BPD, CTR. BPD < CTR. |
| *Clinical* |  |  |  |  |  |
| Age at onset, years (M, SD) | 23.4 (7.6) | 17.9 (5.9) |  | n.a. |  |
| Duration of illness, years (M, SD) | 7.2 (8.5) | 13.1 (8.9) |  | n.a. |  |
| PANSS total score (M, SD) | 59.2 (14.6) | 44.0 (8.0) | | $t=7.6$, $p<0.001$ | |
| Positive factor | 8.6 (3.6) | 5.1 (1.7) | | $t=8.0$, $p<0.001$ | |
| Negative factor | 13.3 (5.2) | 8.5 (3.0) | | $t=7.2$, $p<0.001$ | |
| Disorganized factor | 5.4 (2.4) | 4.0 (1.3) | | $t=4.8$, $p<0.001$ | |
| Excited factor | 5.3 (1.7) | 5.0 (1.3) | | $t=1.4$, $p=0.17$ | |
| Depressive factor | 7.8 (2.8) | 7.9 (2.7) | | $t= -0.25$, $p=0.8$ | |
| GAF – symptoms | 50.4 (11.6) | 62.4 (9.6) | | $t= -7.0$, $p<0.001$ | |
| GAF – functioning | 49.3 (11.8) | 63.8 (11.0) | | $t= -7.2$, $p<0.001$ | |
| *Medication use* |  |  |  |  |  |
| Antipsychotic agents (n, %) | 56 (89%) | 31 (34%) | | $\chi^2 = 45.4$, $p<0.001$ | |
| SGA / FGA / both (n) | 53 / 0 / 3 | 30 / 0 / 1 |  | n.a. |  |
| Oral / depot / both (n) | 41 / 7 / 8 | 30 / 1 / 0 | | $\chi^2 = 7.62$, $p=0.01^1$ | |
| Antipsychotic monotherapy<sup>2</sup> | 39 (70%) | 29 (94%) | | $\chi^2 = 6.68$, $p=0.01^1$ | |
| CPZ-equivalent dose (M, SD) | 318 (158) | 191 (115) | | $t=3.9$, $p<0.001$ | |
| SGA (M, SD) | 305 (162) | 190 (113) | | $t=3.5$, $p<0.001$ | |
| FGA (M, SD) | 254 (185) | 30 (n.a.) |  | n.a. |  |
| Lithium (n, %) | 3 (5%) | 11 (12%) | | $\chi^2 = 2.4$, n.s. | |
| Antiepileptic agents (n, %) | 8 (13%) | 25 (28%) | | $\chi^2 = 4.8$, $p=0.03$ | |
| Antidepressive agents (n, %) | 16 (25%) | 25 (28%) | | $\chi^2 = 0.1$, n.s. | |
| Anxiolytics/hypnotic agents (n, %) | 3 (5%) | 7 (8%) | | $\chi^2 = 0.5$, n.s. | |

Abbreviations: M: Mean. SD: Standard deviation. SCZ: Schizophrenia. BPD: Bipolar disorders. CTR: Healthy controls. WASI: Wechsler Abbreviated Scale of Intelligence. PANSS: Positive and negative syndrome scale. GAF: Global assessment of functioning scale. CPZ: Chlorpromazine. SGA: Second generation antipsychotics. FGA: First-generation antipsychotics. n.s.: Not significant. n.a.: Not applicable.
<sup>1</sup>Calculated using Fisher's exact test.
<sup>2</sup>Defined as the current use of one antipsychotic medication type.
Missing values: Education: n=12. WASI IQ: n=14. Age at onset: n=2. Duration of illness: n=2. GAF – functioning: n=1. PANSS total score: n=1. PANSS negative factor score: n=1. Medication use: n=1. CPZ-equivalent dose: n=1.

### 3.2 Medication use in the patient sample

The use of antipsychotics was more prevalent in SCZ than in BPD, whereas BPD had a more frequent use of antiepileptic drugs. Other medication categories did not differ between groups (Table 1).

Patients with SCZ used higher doses and were more often treated with multiple antipsychotic agents or long-acting injectables than patients with BPD (Table 1). Further information is found in Supplementary Table 1.

The association between age and current antipsychotic dose (CPZ) was not significant (ρ = 0.18, p = 0.09).

### 3.3 No group differences in regional T1w/T2w-ratio values

When examining the main effects of diagnosis (SCZ versus CTR, BPD versus CTR), we found no significant differences in regional T1w/T2w-ratio values between either of the patient groups and CTR.

### 3.4 Different age trajectories of regional T1w/T2w-ratio values

In the group-wise comparison of T1w/T2w-ratio age slopes (i.e., diagnosis x age interaction terms), patients with SCZ had more positive age slopes compared to CTR in 22 clusters. These included clusters in frontal and temporal regions, e.g., bilateral regions of the superior frontal and insular cortices, as well as parietal and occipital regions. There were no significant differences in age slopes between patients with BPD and CTR. An overview of significant clusters is shown in Table 2 and Figure 1. See Supplementary Figure 1 for further details.

**Figure 1.**
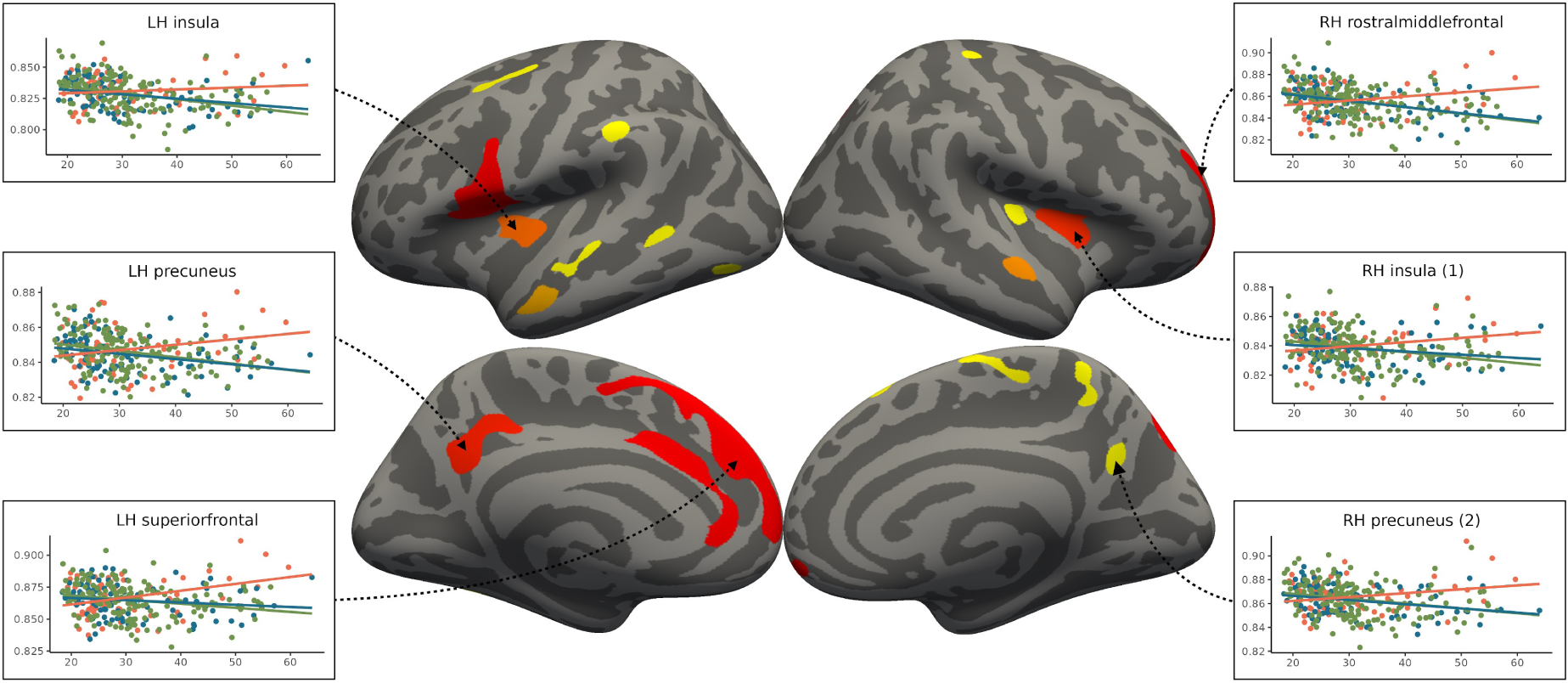
Clusters shown in red to yellow colors on the inflated cortical surfaces signify cortical regions where associations with age differed between SCZ and CTR. The cluster-forming threshold was 0.001 and the cluster-wise probability set at p<0.05 with correction for two hemispheres. In the left and right boxes, scatterplots illustrate the association with age among SCZ (red line), BPD (blue line) and CTR (green line) with age on the x-axes and mean T1w/T2-ratio values within the cluster on the y-axes.

**Table 2.** Significant clusters based on vertex-wise analysis of schizophrenia × age interaction effect

| Region | Maximal<br>significance<br>value | Cluster size<br>(mm<sup>2</sup>) | Coordinates |  |  |
| --- | --- | --- | --- | --- | --- |
|  |  |  | x | y | z |
| *Left hemisphere* |  |  |  |  |  |
| Superior frontal | 1.98 x 10<sup>-6</sup> | 1744 | -8.1 | 44.3 | 35.4 |
| Pars opercularis | 1.56 x 10<sup>-6</sup> | 1239 | -52.0 | 12.9 | 7.6 |
| Caudal anterior cingulate | 7.29 x 10<sup>-6</sup> | 532 | -7.3 | 18.9 | 29.5 |
| Precuneus | 6.49 x 10<sup>-5</sup> | 507 | -12.2 | -55.1 | 29.2 |
| Insula | 8.17 x 10<sup>-5</sup> | 349 | -35.9 | -20.9 | -4.1 |
| Middle temporal | 7.41 x 10<sup>-5</sup> | 249 | -53.8 | -9.0 | -23.1 |
| Precentral | 2.72 x 10<sup>-4</sup> | 229 | -27.5 | -11.1 | 47.0 |
| Superior temporal | 3.00 x 10<sup>-4</sup> | 210 | -54.0 | -23.9 | -3.7 |
| Fusiform gyrus | 2.51 x 10<sup>-4</sup> | 156 | -38.5 | -51.4 | -20.0 |
| Supramarginal | 1.53 x 10<sup>-5</sup> | 139 | -60.4 | -32.5 | 36.3 |
| Lateral occipital | 4.43 x 10<sup>-4</sup> | 125 | -40.5 | -70.9 | -8.1 |
| Banks of the superior temporal sulcus | 1.88 x 10<sup>-4</sup> | 116 | -56.0 | -48.7 | 1.0 |
| *Right hemisphere* |  |  |  |  |  |
| Rostral middle frontal | 4.66 x 10<sup>-6</sup> | 1239 | 18.6 | 56.0 | -5.3 |
| Superior parietal | 1.22 x 10<sup>-4</sup> | 428 | 16.7 | -72.3 | 45.2 |
| Insula (1) | 9.11 x 10<sup>-5</sup> | 364 | 34.0 | 5.7 | 5.9 |
| Superior temporal | 7.91 x 10<sup>-5</sup> | 220 | 57.4 | -8.0 | -7.7 |
| Superior frontal (1) | 2.25 x 10<sup>-4</sup> | 205 | 9.6 | -7.1 | 67.6 |
| Precuneus (1) | 2.47 x 10<sup>-4</sup> | 167 | 10.5 | -48.4 | 61.9 |
| Precuneus (2) | 3.84 x 10<sup>-4</sup> | 130 | 5.2 | -58.9 | 27.3 |
| Insula (2) | 1.04 x 10<sup>-4</sup> | 124 | 34.0 | -25.5 | 6.5 |
| Superior frontal (2) | 4.93 x 10<sup>-4</sup> | 105 | 10.5 | 23.3 | 56.6 |
| Postcentral | 3.16 x 10<sup>-5</sup> | 100 | 27.9 | -30.3 | 58.9 |

Maximal significance value refers to the p-value at the vertex showing the largest difference in age slopes between SCZ and CTR within each cluster.

Follow-up analyses of the linear age slopes in each group showed that CTR had predominantly negative age slopes in medial frontal and temporal regions, with positive age slopes only in the central sulcus. In contrast, patients with SCZ had several regions with positive age slopes, including frontal lobe regions (Figure 2).

**Figure 2.**
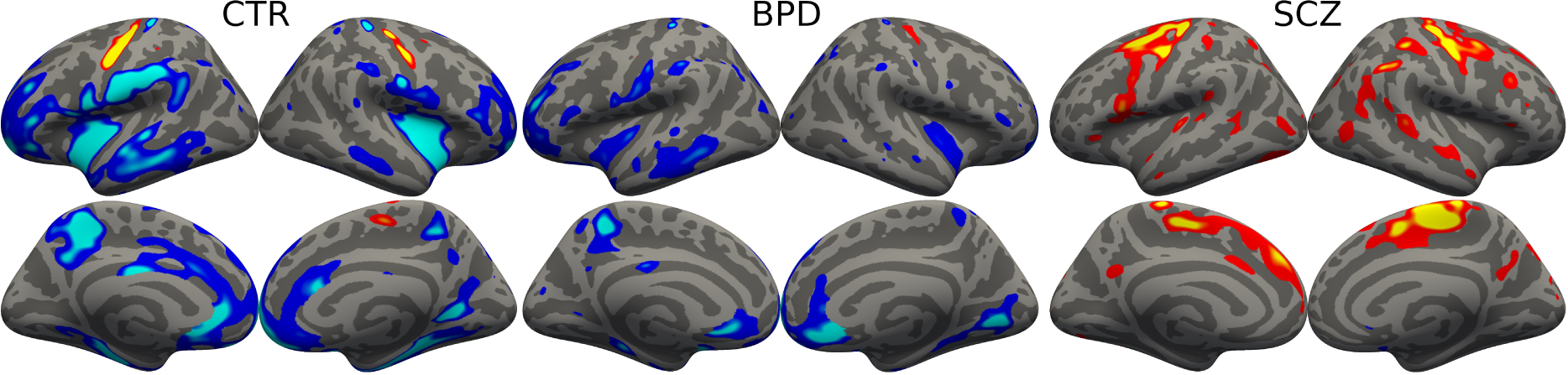
The figure displays results from a vertex-wise general linear model adjusted for age and sex. Contrasts were specified to examine age slopes within each group separately. Colored regions indicate where the association with age deviated from the null hypothesis (i.e., no association) at the threshold p<0.01, uncorrected. Blue to light blue colors denote negative age slopes (i.e., lower T1w/T2w-ratio in older individuals). Red to yellow colors denote positive age slopes (i.e., higher T1w/T2w-ratio values in older individuals).

### 3.5 Age trajectories of the T1w/T2w-ratio and antipsychotic medication use

We found significant interaction effects indicating a more positive age slope in patients using antipsychotic medication compared to patients who did not. This was found in temporal lobe regions, insular regions, the precuneus bilaterally, the left precentral gyrus, the right superior and middle frontal lobe, and the postcentral gyrus (Table 3).

**Table 3.** Analysis of cluster-wise T1w/T2w values, antipsychotic medication use and interaction with age.

| Region | AP |  |  | AP × age |  |  | CPZ |  |  | CPZ × age |  |  |
| --- | --- | --- | --- | --- | --- | --- | --- | --- | --- | --- | --- | --- |
| | $\beta$ | $p$ | $p_{corr}$ | $\beta$ | $p$ | $p_{corr}$ | $\beta$ | $p$ | $p_{corr}$ | $\beta$ | $p$ | $p_{corr}$ |
| *Left hemisphere* |  |  |  |  |  |  |  |  |  |  |  |  |
| Superior frontal | .00 | .99 | .99 | .30 | .02 | .06 | -.04 | .71 | .89 | .32 | .002 | **.02** |
| Pars opercularis | .00 | .97 | .99 | .29 | .02 | .06 | -.12 | .30 | .51 | .26 | .02 | **.05** |
| Caudal anterior cingulate | .04 | .66 | .86 | .27 | .04 | .10 | .00 | .97 | .99 | .26 | .02 | **.05** |
| Precuneus | .07 | .70 | .73 | .36 | .006 | **.03** | -.06 | .63 | .85 | .27 | .01 | **.04** |
| Insula | -.05 | .62 | .85 | .32 | .01 | **.05** | .03 | .77 | .90 | .26 | .02 | **.05** |
| Middle temporal | .04 | .64 | .85 | .33 | .01 | **.04** | -.09 | .45 | .70 | .25 | .02 | **.05** |
| Precentral | -.03 | .75 | .90 | .37 | .004 | **.03** | .08 | .43 | .68 | .33 | .0007 | **.02** |
| Superior temporal | .05 | .63 | .85 | .29 | .03 | .06 | -.13 | .27 | .48 | .31 | .005 | **.03** |
| Fusiform gyrus | -.04 | .67 | .86 | .24 | .08 | .15 | -.21 | .06 | .13 | .28 | .007 | **.03** |
| Supramarginal | .00 | .98 | .99 | .31 | .02 | **.05** | -.08 | .46 | .71 | .32 | .003 | **.02** |
| Lateral occipital | -.04 | .71 | .89 | .28 | .03 | .07 | -.21 | .05 | .12 | .29 | .004 | **.03** |
| Banks of the superior temporal sulcus | .04 | .70 | .89 | .43 | .001 | **.02** | -.17 | .12 | .23 | .33 | .002 | **.02** |
| *Right hemisphere* |  |  |  |  |  |  |  |  |  |  |  |  |
| Rostral middle frontal | -.03 | .76 | .90 | .37 | .005 | **.03** | -.01 | .95 | .99 | .31 | .005 | **.03** |
| Superior parietal | -.02 | .83 | .94 | .21 | .12 | .23 | -.03 | .80 | .92 | .38 | .0004 | **.02** |
| Insula (1) | -.08 | .40 | .64 | .31 | .02 | .05 | -.04 | .73 | .89 | .24 | .03 | .06 |
| Superior temporal | -.02 | .87 | .95 | .34 | .01 | **.04** | -.07 | .54 | .80 | .33 | .002 | **.02** |
| Superior frontal (1) | -.02 | .85 | .95 | .42 | .001 | **.02** | -.12 | .29 | .50 | .27 | .01 | **.04** |
| Precuneus (1) | .07 | .45 | .70 | .39 | .003 | **.02** | .02 | .88 | .96 | .27 | .01 | **.04** |
| Precuneus (2) | .05 | .62 | .85 | .40 | .002 | **.02** | -.15 | .19 | .34 | .33 | .002 | **.02** |
| Insula (2) | .00 | .97 | .99 | .34 | .009 | **.04** | .02 | .86 | .95 | .19 | .08 | .16 |
| Superior frontal (2) | .01 | .95 | .99 | .48 | .0001 | **.01** | .06 | .57 | .82 | .23 | .01 | **.04** |
| Postcentral | .14 | .16 | .29 | .32 | .01 | **.04** | -.10 | .37 | .62 | .20 | .06 | .12 |

Linear regression models (n=154) adjusted for age, sex and diagnosis. The age and CPZ variables were centered prior to analysis. P-values below 0.05 (FDR-corrected) are marked in bold. Information about antipsychotic medication use was missing for one participant.
Abbreviations: AP = Antipsychotic medication status. CPZ = Chlorpromazine-equivalent dose.

When analyses were further restricted to patients using antipsychotic medication only (n=86), we found more positive age slopes of T1w/T2w-ratio values in patients using a higher current dose (CPZ). Significant CPZ × age interaction effects were found in all except two of the 22 significant clusters (Table 3).

### 3.6 No association with clinical symptoms

We found no associations between T1w/T2w-ratio values and PANSS total scores, nor with any of the five symptom factors after correction for multiple testing. See Supplementary Table 2 for details.

## 4. Discussion

We did not find lower T1w/T2w-ratio values in either of the patient groups compared to healthy controls. Thus, insofar as the T1w/T2w-ratio is a measure of intracortical myelin, our results provide little support for intracortical myelin deficits in these disorders. However, we observed divergent age trajectories in patients with schizophrenia spectrum disorders. Antipsychotic medication status and dose were both associated with divergent age slopes within the patient sample, which is consistent with a possible promyelinating effect of antipsychotic medication.

The absence of lower regional T1w/T2w-ratio values in patients contrasts with our previous study on the GWC (Jorgensen et al., 2016). While these are different measures, they show moderate to high correlations, with a reported overall correlation of 0.73 (Parent et al., 2023). Our results also differed from previous findings of lower global and regional T1w/T2w-ratio values in patients with schizophrenia spectrum disorders (Ganzetti et al., 2015; Iwatani et al., 2015). However, it is worth noting that only the study by Ganzetti et al (2015) reported lower *regional* T1w/T2w-ratio values. In this study, data was pooled from three different sites, which may have influenced the results given the known effects of scanner on the T1w/T2w-ratio (Nerland et al., 2021). Furthermore, the calibration method based on small masks covering the eyes and the temporal muscles may be unreliable, especially for low-resolution data. In the study by Iwatani et al (2015), a large smoothing kernel was used for computing the T1w/T2w-ratio, and a large portion of the sensorimotor cortices was excluded from the analyses. Notably, both these previous studies used low-resolution T2w images, which may introduce partial volume effects (Shafee et al., 2015).

In two recent studies on first-episode treatment-naïve patients with schizophrenia spectrum disorders (FEP) by Wei and colleagues (2022; 2020), a layer-dependent regional pattern was reported, with lower T1w/T2w-ratio values in the left cingulate and insula and higher values in the left superior temporal gyrus. Notably, these regions belong to the salience network and the language and auditory processing circuitry, respectively, both thought to be affected in psychotic disorders. Interestingly, the patients with FEP differed from healthy controls in the superficial and middle layers of the cortex, but not in the deep layer. In the present study, T1w/T2w-ratio values were sampled at distances of 10-80% of cortical thickness from the gray-white surface, and depth-dependent analyses were not performed. Furthermore, Wei et al (2020) normalized each individual T1w/T2w-ratio map by subtracting the subject-wise mean T1w/T2w-ratio and dividing by the variance, which makes the comparison to the present study difficult.

We observed more positive age trajectories of the T1w/T2w-ratio in patients with schizophrenia spectrum disorders. Furthermore, patients currently treated with antipsychotics showed more positive age trajectories compared with patients not currently on antipsychotics, and there was evidence of a dose-response relationship (higher CPZ associated with a more positive age slope). It has been proposed that some antipsychotic medications have myelin-promoting properties (Bartzokis, 2012) and that their lipogenic effects are related to clinical efficacy (Kim et al., 2019; Leucht et al., 2013; Procyshyn et al., 2007). Indeed, many psychotropic drugs are known to induce cholesterol synthesis (Ferno et al., 2011), and there is evidence to suggest that similar effects are present in human settings (Barth et al., 2020; Bartzokis et al., 2007; Gjerde et al., 2018b; Tishler et al., 2018). Thus, the interpretation that divergent age trajectories in patients reflect antipsychotic treatment is plausible. To directly test this hypothesis, we encourage future studies to assess intracortical myelination longitudinally in patients with FEP who are drug-naïve at baseline.

### 4.1 Strengths and limitations

Strengths of this study include the use of a clinically well-characterized sample where all participants were scanned on the same MRI system. We used a well-tested pipeline for computing the T1w/T2w-ratio, which showed good test-retest reliability and agreement with known myeloarchitecture using scan acquisitions from the same MRI scanner system and pulse sequence parameters.

To compute the T1w/T2w-ratio, we used an intensity normalization procedure, WhiteStripe, based on intensities in normal-appearing white matter (NAWM). While this was previously shown to improve test-retest reliability whilst preserving individual variation in T1w/T2w-ratio distributions, it may introduce dependencies between cortical T1w/T2w-ratio values and T1w and T2w intensity values in NAWM. We cannot rule out the possibility that the observed age-by-diagnosis interactions reflect age trajectories of NAWM rather than gray matter. Quantitative MRI pulse sequences, such as inversion recovery imaging, may be used to rule out this possibility. Such methods estimate biophysically meaningful properties of the MRI measurements and can be used for between-subject comparisons without the need for intensity normalization or calibration.

The T1w/T2w-ratio shows spatial correlation with cortical myeloarchitecture (Glasser et al., 2014) but is based on T1w and T2w image intensities which are inherently non-dimensional measures. Recent studies have indicated that the correlations between the T1w/T2w-ratio and other indices of myelination vary between brain regions. For instance, low correlations have been found with the myelin-water fraction (MWF) in densely myelinated regions in white matter (Sandrone et al., 2023; Uddin et al., 2019). High correlations have, however, been found between the T1w/T2w-ratio and T1 relaxation time mapping of the cerebral cortex (Parent et al., 2023; Shams et al., 2019). Still, strong conclusions regarding microstructural tissue properties should be avoided since the T1w/T2w-ratio remains a complex measure and other tissue properties than myelin content, such as iron content or dendritic density, may also influence it (Righart et al., 2017).

Since we employed a cross-sectional and naturalistic study design, it was not possible to identify causal mechanisms or investigate longitudinal T1w/T2w-ratio change. Although our study sample was large, given the clinical context, it may have been underpowered to detect subtle alterations in the T1w/T2w-ratio. The T1w and T2w sequences we used both had 1mm isotropic resolution; however, for cortical myelin mapping, images with submillimeter resolution would be optimal. Furthermore, while our optimized pipeline improved reliability (Nerland et al., 2021), correction for field inhomogeneities through the acquisition of B1+ field maps is an alternative approach (Glasser et al., 2022). Lastly, although our findings did not confirm an intracortical myelin deficit in schizophrenia spectrum disorders, such deficits could be present in early illness phases or in treatment-naïve individuals.

### 4.2 Conclusions

While our findings did not support the hypothesis of intracortical myelin deficits in schizophrenia spectrum or bipolar disorders, they were consistent with the hypothesized promyelinating effect of antipsychotic medication. The possibility that this effect could also mask an intracortical myelin deficit in patients cannot be ruled out. The findings should be followed up by applying quantitative MRI measures and assessing longitudinal trajectories of intracortical myelination in patients who are drug-naïve at baseline.

## 5. Conflict of interest

OAA has received a speaker’s honorarium from Lundbeck and Sunovion and is a consultant for HealthLytix. IA has received a speaker’s honorarium from Lundbeck. The other authors report no conflict of interest.

## Data Availability

The data was collected by the Norwegian Research Centre for Mental Disorders (NORMENT) at the Oslo University Hospital (OUS). The data is subject to restrictions and is not publicly available but may be made available given reasonable request. Data can only be made available following permission from OUS, and insofar requests are in line with the relevant ethical agreements and the consent of the participants.

## Acknowledgements

We thank the study participants and the clinicians responsible for recruitment and assessment at the Norwegian Research Centre for Mental Disorders (NORMENT). We also thank the scientific assistants who performed quality assurance and editing of reconstructed surfaces. The work was partly conducted on a platform provided by the Services for sensitive data (TSD), operated and developed at the University of Oslo IT Department (USIT).

## 6. Author contributions

Authors IA and OAA took part in designing the project and data acquisition. Authors KNJ and SN were responsible for the MRI post-processing and quality control. Authors KNJ and SN undertook the statistical analyses. All authors contributed to the interpretation of results. Authors IA, EGJ, OMG and IIM supervised the study. Authors KNJ and SN wrote the initial draft of the manuscript. All authors critically reviewed and provided input towards the final version. All authors have contributed to and approved the manuscript.

## 7. Role of the funding source

This work was supported by The Research Council of Norway (grant numbers 223273, 274359) and the South-Eastern Norway Regional Health Authority (grant number 2019-104). The funding sources had no further role in study design, data collection, analysis and interpretation of data, writing the manuscript, or submitting the paper for publication.

## Notes

### Author Declarations

The study complied with the Helsinki Declaration and was approved by the Regional Committee for Medical Research Ethics (REC South-East Norway) and the Norwegian Data Inspectorate. All participants gave informed consent.

## References

Aminoff, S.R., Onyeka, I.N., Ødegaard, M., Simonsen, C., Lagerberg, T.V., Andreassen, O.A., Romm, K.L., Melle, I., 2022. Lifetime and point prevalence of psychotic symptoms in adults with bipolar disorders: a systematic review and meta-analysis. Psychological medicine 52(13), 2413–2425.

Andreasen, N.C., Pressler, M., Nopoulos, P., Miller, D., Ho, B.C., 2010. Antipsychotic dose equivalents and dose-years: a standardized method for comparing exposure to different drugs. Biological psychiatry 67(3), 255–262.

Barth, C., Lonning, V., Gurholt, T.P., Andreassen, O.A., Myhre, A.M., Agartz, I., 2020. Exploring white matter microstructure and the impact of antipsychotics in adolescent-onset psychosis. PLoS One 15(5), e0233684.

Bartzokis, G., 2002. Schizophrenia: breakdown in the well-regulated lifelong process of brain development and maturation. Neuropsychopharmacology 27(4), 672–683.

Bartzokis, G., 2012. Neuroglialpharmacology: myelination as a shared mechanism of action of psychotropic treatments. Neuropharmacology 62(7), 2137–2153.

Bartzokis, G., Lu, P.H., Nuechterlein, K.H., Gitlin, M., Doi, C., Edwards, N., Lieu, C., Altshuler, L.L., Mintz, J., 2007. Differential effects of typical and atypical antipsychotics on brain myelination in schizophrenia. Schizophrenia research 93(1-3), 13–22.

Beer, A., Biberacher, V., Schmidt, P., Righart, R., Buck, D., Berthele, A., Kirschke, J., Zimmer, C., Hemmer, B., Muhlau, M., 2016. Tissue damage within normal appearing white matter in early multiple sclerosis: assessment by the ratio of T1- and T2-weighted MR image intensity. Journal of neurology 263(8), 1495–1502.

Ersland, K.M., Skrede, S., Stansberg, C., Steen, V.M., 2017. Subchronic olanzapine exposure leads to increased expression of myelination-related genes in rat fronto-medial cortex. Translational psychiatry 7(11), 1262.

Ferno, J., Skrede, S., Vik-Mo, A.O., Jassim, G., Le Hellard, S., Steen, V.M., 2011. Lipogenic effects of psychotropic drugs: focus on the SREBP system. Frontiers in bioscience (Landmark edition) 16, 49–60.

Fischl, B., 2012. FreeSurfer. NeuroImage 62(2), 774–781.

Friston, K., Brown, H.R., Siemerkus, J., Stephan, K.E., 2016. The dysconnection hypothesis (2016). Schizophrenia research 176(2-3), 83–94.

Friston, K.J., Frith, C.D., 1995. Schizophrenia: a disconnection syndrome? Clinical neuroscience (New York, N.Y.) 3(2), 89–97.

Ganzetti, M., Wenderoth, N., Mantini, D., 2015. Mapping pathological changes in brain structure by combining T1- and T2-weighted MR imaging data. Neuroradiology 57(9), 917–928.

Gjerde, P.B., Dieset, I., Simonsen, C., Hoseth, E.Z., Iversen, T., Lagerberg, T.V., Lyngstad, S.H., Mørch, R.H., Skrede, S., Andreassen, O.A., Melle, I., Steen, V.M., 2018a. Increase in serum HDL level is associated with less negative symptoms after one year of antipsychotic treatment in first-episode psychosis. Schizophrenia research 197, 253–260.

Gjerde, P.B., Jørgensen, K.N., Steen, N.E., Melle, I., Andreassen, O.A., Steen, V.M., Agartz, I., 2018b. Association between olanzapine treatment and brain cortical thickness and gray/white matter contrast is moderated by cholesterol in psychotic disorders. Psychiatry Research: Neuroimaging 282, 55–63.

Glasser, M.F., Coalson, T.S., Harms, M.P., Xu, J., Baum, G.L., Autio, J.A., Auerbach, E.J., Greve, D.N., Yacoub, E., Van Essen, D.C., Bock, N.A., Hayashi, T., 2022. Empirical transmit field bias correction of T1w/T2w myelin maps. NeuroImage 258, 119360.

Glasser, M.F., Coalson, T.S., Robinson, E.C., Hacker, C.D., Harwell, J., Yacoub, E., Ugurbil, K., Andersson, J., Beckmann, C.F., Jenkinson, M., Smith, S.M., Van Essen, D.C., 2016. A multi-modal parcellation of human cerebral cortex. Nature 536(7615), 171–178.

Glasser, M.F., Goyal, M.S., Preuss, T.M., Raichle, M.E., Van Essen, D.C., 2014. Trends and properties of human cerebral cortex: correlations with cortical myelin content. NeuroImage 93 Pt 2, 165–175.

Glasser, M.F., Van Essen, D.C., 2011. Mapping human cortical areas in vivo based on myelin content as revealed by T1- and T2-weighted MRI. The Journal of neuroscience: the official journal of the Society for Neuroscience 31(32), 11597–11616.

Goudriaan, A., de Leeuw, C., Ripke, S., Hultman, C.M., Sklar, P., Sullivan, P.F., Smit, A.B., Posthuma, D., Verheijen, M.H., 2014. Specific glial functions contribute to schizophrenia susceptibility. Schizophrenia bulletin 40(4), 925–935.

Greve, D.N., Fischl, B., 2009. Accurate and robust brain image alignment using boundary-based registration. NeuroImage 48(1), 63–72.

Hakak, Y., Walker, J.R., Li, C., Wong, W.H., Davis, K.L., Buxbaum, J.D., Haroutunian, V., Fienberg, A.A., 2001. Genome-wide expression analysis reveals dysregulation of myelination-related genes in chronic schizophrenia. Proceedings of the National Academy of Sciences of the United States of America 98(8), 4746–4751.

Ho, D., Imai, K., King, G., Stuart, E.A., 2011. MatchIt: Nonparametric Preprocessing for Parametric Causal Inference. Journal of Statistical Software 42(8), 1–28.

Ishida, T., Donishi, T., Iwatani, J., Yamada, S., Takahashi, S., Ukai, S., Shinosaki, K., Terada, M., Kaneoke, Y., 2017. Elucidating the aberrant brain regions in bipolar disorder using T1-weighted/T2-weighted magnetic resonance ratio images. Psychiatry research. Neuroimaging 263, 76–84.

Iwatani, J., Ishida, T., Donishi, T., Ukai, S., Shinosaki, K., Terada, M., Kaneoke, Y., 2015. Use of T1-weighted/T2-weighted magnetic resonance ratio images to elucidate changes in the schizophrenic brain. Brain and behavior 5(10), e00399.

Jorgensen, K.N., Nerland, S., Norbom, L.B., Doan, N.T., Nesvag, R., Morch-Johnsen, L., Haukvik, U.K., Melle, I., Andreassen, O.A., Westlye, L.T., Agartz, I., 2016. Increased MRI-based cortical grey/white-matter contrast in sensory and motor regions in schizophrenia and bipolar disorder. Psychological medicine, 1-15.

Kay, S.R., Fiszbein, A., Opler, L.A., 1987. The positive and negative syndrome scale (PANSS) for schizophrenia. Schizophrenia bulletin 13(2), 261–276.

Kelly, S., Jahanshad, N., Zalesky, A., Kochunov, P., Agartz, I., Alloza, C., Andreassen, O.A., Arango, C., Banaj, N., Bouix, S., Bousman, C.A., Brouwer, R.M., Bruggemann, J., Bustillo, J., Cahn, W., Calhoun, V., Cannon, D., Carr, V., Catts, S., Chen, J., Chen, J.X., Chen, X., Chiapponi, C., Cho, K.K., Ciullo, V., Corvin, A.S., Crespo-Facorro, B., Cropley, V., De Rossi, P., Diaz-Caneja, C.M., Dickie, E.W., Ehrlich, S., Fan, F.M., Faskowitz, J., Fatouros-Bergman, H., Flyckt, L., Ford, J.M., Fouche, J.P., Fukunaga, M., Gill, M., Glahn, D.C., Gollub, R., Goudzwaard, E.D., Guo, H., Gur, R.E., Gur, R.C., Gurholt, T.P., Hashimoto, R., Hatton, S.N., Henskens, F.A., Hibar, D.P., Hickie, I.B., Hong, L.E., Horacek, J., Howells, F.M., Hulshoff Pol, H.E., Hyde, C.L., Isaev, D., Jablensky, A., Jansen, P.R., Janssen, J., Jönsson, E.G., Jung, L.A., Kahn, R.S., Kikinis, Z., Liu, K., Klauser, P., Knöchel, C., Kubicki, M., Lagopoulos, J., Langen, C., Lawrie, S., Lenroot, R.K., Lim, K.O., Lopez-Jaramillo, C., Lyall, A., Magnotta, V., Mandl, R.C.W., Mathalon, D.H., McCarley, R.W., McCarthy-Jones, S., McDonald, C., McEwen, S., McIntosh, A., Melicher, T., Mesholam-Gately, R.I., Michie, P.T., Mowry, B., Mueller, B.A., Newell, D.T., O’Donnell, P., Oertel-Knöchel, V., Oestreich, L., Paciga, S.A., Pantelis, C., Pasternak, O., Pearlson, G., Pellicano, G.R., Pereira, A., Pineda Zapata, J., Piras, F., Potkin, S.G., Preda, A., Rasser, P.E., Roalf, D.R., Roiz, R., Roos, A., Rotenberg, D., Satterthwaite, T.D., Savadjiev, P., Schall, U., Scott, R.J., Seal, M.L., Seidman, L.J., Shannon Weickert, C., Whelan, C.D., Shenton, M.E., Kwon, J.S., Spalletta, G., Spaniel, F., Sprooten, E., Stäblein, M., Stein, D.J., Sundram, S., Tan, Y., Tan, S., Tang, S., Temmingh, H.S., Westlye, L.T., Tønnesen, S., Tordesillas-Gutierrez, D., Doan, N.T., Vaidya, J., van Haren, N.E.M., Vargas, C.D., Vecchio, D., Velakoulis, D., Voineskos, A., Voyvodic, J.Q., Wang, Z., Wan, P., Wei, D., Weickert, T.W., Whalley, H., White, T., Whitford, T.J., Wojcik, J.D., Xiang, H., Xie, Z., Yamamori, H., Yang, F., Yao, N., Zhang, G., Zhao, J., van Erp, T.G.M., Turner, J., Thompson, P.M., Donohoe, G., 2018. Widespread white matter microstructural differences in schizophrenia across 4322 individuals: results from the ENIGMA Schizophrenia DTI Working Group. Molecular psychiatry 23(5), 1261–1269.

Kim, D.D., Barr, A.M., Fredrikson, D.H., Honer, W.G., Procyshyn, R.M., 2019. Association between Serum Lipids and Antipsychotic Response in Schizophrenia. Curr Neuropharmacol 17(9), 852–860.

Koenig, S.H., 1991. Cholesterol of myelin is the determinant of gray-white contrast in MRI of brain. Magn Reson Med 20(2), 285–291.

Koenig, S.H., Brown, R.D., 3rd, Spiller, M., Lundbom, N., 1990. Relaxometry of brain: why white matter appears bright in MRI. Magn Reson Med 14(3), 482-495.

Kolomeets, N.S., Uranova, N.A., 2018. Reduced oligodendrocyte density in layer 5 of the prefrontal cortex in schizophrenia. European archives of psychiatry and clinical neuroscience.

Kroken, R.A., Løberg, E.-M., Drønen, T., Gruner, R., Hugdahl, K., Kompus, K., Skrede, S., Johnsen, E., 2014. A Critical Review of Pro-Cognitive Drug Targets in Psychosis: Convergence on Myelination and Inflammation. Frontiers in Psychiatry 5.

Leucht, S., Cipriani, A., Spineli, L., Mavridis, D., Orey, D., Richter, F., Samara, M., Barbui, C., Engel, R.R., Geddes, J.R., Kissling, W., Stapf, M.P., Lassig, B., Salanti, G., Davis, J.M., 2013. Comparative efficacy and tolerability of 15 antipsychotic drugs in schizophrenia: a multiple-treatments meta-analysis. Lancet 382(9896), 951–962.

Makowski, C., Lewis, J.D., Lepage, C., Malla, A.K., Joober, R., Lepage, M., Evans, A.C., 2019. Structural Associations of Cortical Contrast and Thickness in First Episode Psychosis. Cerebral cortex (New York, N.Y.: 1991).

Necus, J., Smith, F.E., Thelwall, P.E., Flowers, C.J., Sinha, N., Taylor, P.N., Blamire, A.M., Wang, Y., Cousins, D.A., 2019. Quantification of brain proton longitudinal relaxation (T1) in lithium-treated and lithium-naive patients with bipolar disorder in comparison to healthy controls. Bipolar disorders.

Nerland, S., Jørgensen, K.N., Nordhøy, W., Maximov, II, Bugge, R.A.B., Westlye, L.T., Andreassen, O.A., Geier, O.M., Agartz, I., 2021. Multisite reproducibility and test-retest reliability of the T1w/T2w-ratio: A comparison of processing methods. NeuroImage 245, 118709.

Parent, O., Olafson, E., Bussy, A., Tullo, S., Blostein, N., Dai, A., Salaciak, A., Bedford, S.A., Farzin, S., Béland, M.L., Valiquette, V., Tardif, C.L., Devenyi, G.A., Chakravarty, M.M., 2023. High spatial overlap but diverging age-related trajectories of cortical magnetic resonance imaging markers aiming to represent intracortical myelin and microstructure. Human brain mapping 44(8), 3023–3044.

Paus, T., Keshavan, M., Giedd, J.N., 2008. Why do many psychiatric disorders emerge during adolescence? Nature reviews. Neuroscience 9(12), 947–957.

Pearlson, G.D., 2015. Etiologic, phenomenologic, and endophenotypic overlap of schizophrenia and bipolar disorder. Annu Rev Clin Psychol 11, 251–281.

Pedersen, G., Hagtvet, K.A., Karterud, S., 2007. Generalizability studies of the Global Assessment of Functioning-Split version. Compr.Psychiatry 48(1), 88–94.

Pelkmans, W., Dicks, E., Barkhof, F., Vrenken, H., Scheltens, P., van der Flier, W.M., Tijms, B.M., 2019. Gray matter T1-w/T2-w ratios are higher in Alzheimer’s disease. Human brain mapping 40(13), 3900–3909.

Perälä, J., Suvisaari, J., Saarni, S.I., Kuoppasalmi, K., Isometsä, E., Pirkola, S., Partonen, T., Tuulio-Henriksson, A., Hintikka, J., Kieseppä, T., Härkänen, T., Koskinen, S., Lönnqvist, J., 2007. Lifetime prevalence of psychotic and bipolar I disorders in a general population. Arch Gen Psychiatry 64(1), 19–28.

Procyshyn, R.M., Wasan, K.M., Thornton, A.E., Barr, A.M., Chen, E.Y.H., Pomarol-Clotet, E., Stip, E., Williams, R., MacEwan, G.W., Birmingham, C.L., Honer, W.G., for the, C., Risperidone Enhancement Study, G., 2007. Changes in serum lipids, independent of weight, are associated with changes in symptoms during long-term clozapine treatment. Journal of psychiatry & neuroscience: JPN 32(5), 331–338.

Rangel-Guerra, R.A., Perez-Payan, H., Minkoff, L., Todd, L.E., 1983. Nuclear magnetic resonance in bipolar affective disorders. AJNR Am J Neuroradiol 4(3), 229–231.

Righart, R., Biberacher, V., Jonkman, L.E., Klaver, R., Schmidt, P., Buck, D., Berthele, A., Kirschke, J.S., Zimmer, C., Hemmer, B., Geurts, J.J.G., Mühlau, M., 2017. Cortical pathology in multiple sclerosis detected by the T1/T2-weighted ratio from routine magnetic resonance imaging. Ann Neurol 82(4), 519–529.

Rowley, C.D., Tabrizi, S.J., Scahill, R.I., Leavitt, B.R., Roos, R.A.C., Durr, A., Bock, N.A., 2018. Altered Intracortical T1-Weighted/T2-Weighted Ratio Signal in Huntington’s Disease. Frontiers in neuroscience 12, 805.

Sandrone, S., Aiello, M., Cavaliere, C., Thiebaut de Schotten, M., Reimann, K., Troakes, C., Bodi, I., Lacerda, L., Monti, S., Murphy, D., Geyer, S., Catani, M., Dell’Acqua, F., 2023. Mapping myelin in white matter with T1-weighted/T2-weighted maps: discrepancy with histology and other myelin MRI measures. Brain structure & function 228(2), 525–535.

Sehmbi, M., Rowley, C.D., Minuzzi, L., Kapczinski, F., Kwiecien, J.M., Bock, N.A., Frey, B.N., 2019. Age-related deficits in intracortical myelination in young adults with bipolar disorder type I. Journal of psychiatry & neuroscience: JPN 44(2), 79–88.

Sehmbi, M., Rowley, C.D., Minuzzi, L., Kapczinski, F., Steiner, M., Sassi, R.B., Bock, N.A., Frey, B.N., 2018. Association of intracortical myelin and cognitive function in bipolar I disorder. Acta Psychiatr Scand 138(1), 62–72.

Shafee, R., Buckner, R.L., Fischl, B., 2015. Gray matter myelination of 1555 human brains using partial volume corrected MRI images. NeuroImage 105, 473–485.

Shams, Z., Norris, D.G., Marques, J.P., 2019. A comparison of in vivo MRI based cortical myelin mapping using T1w/T2w and R1 mapping at 3T. PLoS One 14(7), e0218089.

Spitzer, R.L., Williams, J.B., Gibbon, M., First, M.B., 1992. The Structured Clinical Interview for DSM-III-R (SCID). I: History, rationale, and description. Archives of General Psychiatry 49(8), 624–629.

Spitzer, R.L., Williams, J.B., Kroenke, K., Linzer, M., deGruy, F.V., 3rd, Hahn, S.R., Brody, D., Johnson, J.G., 1994. Utility of a new procedure for diagnosing mental disorders in primary care. The PRIME-MD 1000 study. Jama 272(22), 1749-1756.

Tamminga, C.A., Pearlson, G., Keshavan, M., Sweeney, J., Clementz, B., Thaker, G., 2014. Bipolar and Schizophrenia Network for Intermediate Phenotypes: Outcomes Across the Psychosis Continuum. Schizophrenia bulletin 40(Suppl_2), S131-S137.

Tishler, T.A., Bartzokis, G., Lu, P.H., Raven, E.P., Khanoyan, M., Kirkpatrick, C.J., Pyle, M.H., Villablanca, J.P., Altshuler, L.L., Mintz, J., Ventura, J., Casaus, L.R., Subotnik, K.L., Nuechterlein, K.H., Ellingson, B.M., 2018. Abnormal Trajectory of Intracortical Myelination in Schizophrenia Implicates White Matter in Disease Pathophysiology and the Therapeutic Mechanism of Action of Antipsychotics. Biological psychiatry. Cognitive neuroscience and neuroimaging 3(5), 454–462.

Uddin, M.N., Figley, T.D., Solar, K.G., Shatil, A.S., Figley, C.R., 2019. Comparisons between multi-component myelin water fraction, T1w/T2w ratio, and diffusion tensor imaging measures in healthy human brain structures. Scientific reports 9(1), 2500.

Uranova, N.A., Vikhreva, O.V., Rachmanova, V.I., Orlovskaya, D.D., 2011. Ultrastructural alterations of myelinated fibers and oligodendrocytes in the prefrontal cortex in schizophrenia: a postmortem morphometric study. Schizophrenia research and treatment 2011, 325789.

van Bergen, A.H., Verkooijen, S., Vreeker, A., Abramovic, L., Hillegers, M.H., Spijker, A.T., Hoencamp, E., Regeer, E.J., Knapen, S.E., Riemersma-van der Lek, R.F., Schoevers, R., Stevens, A.W., Schulte, P.F.J., Vonk, R., Hoekstra, R., van Beveren, N.J., Kupka, R.W., Sommer, I.E.C., Ophoff, R.A., Kahn, R.S., Boks, M.P.M., 2019. The characteristics of psychotic features in bipolar disorder. Psychological medicine 49(12), 2036–2048.

Vikhreva, O.V., Rakhmanova, V.I., Orlovskaya, D.D., Uranova, N.A., 2016. Ultrastructural alterations of oligodendrocytes in prefrontal white matter in schizophrenia: A post-mortem morphometric study. Schizophrenia research.

Waehnert, M.D., Dinse, J., Schäfer, A., Geyer, S., Bazin, P.L., Turner, R., Tardif, C.L., 2016. A subject-specific framework for in vivo myeloarchitectonic analysis using high resolution quantitative MRI. NeuroImage 125, 94–107.

Wallwork, R.S., Fortgang, R., Hashimoto, R., Weinberger, D.R., Dickinson, D., 2012. Searching for a consensus five-factor model of the Positive and Negative Syndrome Scale for schizophrenia. Schizophrenia research 137(1-3), 246–250.

Wechsler, D., 2007. Wechsler Abbreviated Scale of Intelligence (WASI). Norwegian manual supplement. Pearson assessment, Stockholm.

Wei, W., Yin, Y., Zhang, Y., Li, X., Li, M., Guo, W., Wang, Q., Deng, W., Ma, X., Zhao, L., Palaniyappan, L., Li, T., 2022. Structural Covariance of Depth-Dependent Intracortical Myelination in the Human Brain and Its Application to Drug-Naïve Schizophrenia: A T1w/T2w MRI Study. Cerebral cortex (New York, N.Y.: 1991) 32(11), 2373-2384.

Wei, W., Zhang, Y., Li, Y., Meng, Y., Li, M., Wang, Q., Deng, W., Ma, X., Palaniyappan, L., Zhang, N., Li, T., 2020. Depth-dependent abnormal cortical myelination in first-episode treatment-naïve schizophrenia. Human brain mapping 41(10), 2782–2793.

Whitford, T.J., Ford, J.M., Mathalon, D.H., Kubicki, M., Shenton, M.E., 2012. Schizophrenia, myelination, and delayed corollary discharges: a hypothesis. Schizophrenia bulletin 38(3), 486–494.

Xia, M., Womer, F.Y., Chang, M., Zhu, Y., Zhou, Q., Edmiston, E.K., Jiang, X., Wei, S., Duan, J., Xu, K., Tang, Y., He, Y., Wang, F., 2019. Shared and Distinct Functional Architectures of Brain Networks Across Psychiatric Disorders. Schizophrenia bulletin 45(2), 450–463.

