## Supplementary Materials for "Assessing regional intracortical myelination in schizophrenia spectrum and bipolar disorders using the optimized T1w/T2w-ratio"

**Content:**

Supplementary Table 1

Supplementary Table 2

Supplementary Figure 1

Preprint version

**Supplementary Table 1: Distribution of antipsychotic medication types used.**

| | SCZ (n=56) | BPD (n=31) | All patients (n=87) |
|---|---|---|---|
| *SGA* | | | |
| Olanzapine (N05AH03) | 26 (46%) | 4 (13%) | 31 (35%) |
| Aripiprazole (N05AX12) | 17 30%) | 11 (35%) | 28 (32%) |
| Quetiapine (N05AH04) | 5 (9%) | 13 (42%) | 18 (20%) |
| Risperidone (N05AX08) | 9 (16%) | 2 (6%) | 11 (13%) |
| Paliperidone (N05AX13) | 8 (14%) | 1 (3%) | 9 (10%) |
| Ziprazidone (N05AE04) | 3 (5%) | 0 (0%) | 3 (3%) |
| Clozapine (N05AH02) | 2 (3%) | 0 (0%) | 2 (2%) |
| *FGA* | | | |
| Zuclopenthixol (N05AF05) | 2 (3%) | 0 (0%) | 2 (2%) |
| Haloperidol (N05AD01) | 1 (2%) | 0 (0%) | 1 (1%) |
| Chlorprothixene (N05AF03) | 0 (0%) | 1 (3%) | 1 (1%) |

Anatomical Therapeutic Chemical (ATC) classification codes are provided in the parentheses.

Abbreviations: SCZ: Schizophrenia. BPD: Bipolar disorder. SGA: Second generation antipsychotic agent. FGA: First generation antipsychotic agent.

Preprint version

**Supplementary Table 1 – Associations with PANSS total scores and symptom factors.**

| Region | PANSS total score[1] | | | Positive factor | | | Negative factor | | | Disorganized factor | | | Excited factor | | | Depressive factor | | |
|---|---|---|---|---|---|---|---|---|---|---|---|---|---|---|---|---|---|---|
| | *β* | *p* | $p_{corr}$ | *β* | *p* | $p_{corr}$ | *β* | *p* | $p_{corr}$ | *β* | *p* | $p_{corr}$ | *β* | *p* | $p_{corr}$ | *β* | *p* | $p_{corr}$ |
| *Left hemisphere* | | | | | | | | | | | | | | | | | | |
| Superior frontal | .04 | .68 | .96 | .10 | .32 | .94 | .01 | .91 | .98 | .08 | .36 | .94 | .14 | .09 | .76 | -.07 | .37 | .94 |
| Pars opercularis | .03 | .76 | .96 | .05 | .51 | .96 | -.03 | .72 | .96 | .10 | .24 | .83 | .14 | .07 | .76 | -.03 | .70 | .96 |
| Caudal anterior cingulate | .04 | .68 | .96 | .07 | .45 | .96 | .02 | .81 | .97 | .05 | .55 | .96 | .13 | .10 | .76 | -.07 | .41 | .94 |
| Precuneus | .05 | .61 | .96 | .14 | .14 | .76 | -.04 | .67 | .96 | .05 | .55 | .96 | .14 | .09 | .76 | -.04 | .65 | .96 |
| Insula | .02 | .81 | .97 | .12 | .20 | .76 | -.03 | .79 | .97 | .17 | .04 | .76 | .11 | .17 | .76 | -.11 | .18 | .76 |
| Middle temporal | .07 | .45 | .96 | .10 | .32 | .94 | .00 | .99 | .99 | .14 | .10 | .76 | .16 | .05 | .76 | -.01 | .89 | .98 |
| Precentral | .00 | .99 | .99 | .05 | .63 | .96 | -.01 | .91 | .98 | .07 | .42 | .94 | .08 | .30 | .94 | -.03 | .70 | .96 |
| Superior temporal | .05 | .47 | .96 | .09 | .32 | .94 | -.04 | .68 | .96 | .12 | .17 | .76 | .11 | .17 | .76 | -.03 | .70 | .96 |
| Fusiform gyrus | -.03 | .76 | .96 | .09 | .35 | .94 | -.01 | .89 | .98 | -.03 | .71 | .96 | .07 | .40 | .94 | -.05 | .55 | .96 |
| Supramarginal | -.05 | .62 | .96 | .03 | .73 | .96 | -.12 | .20 | .76 | .08 | .36 | .94 | .11 | .20 | .76 | -.03 | .72 | .96 |
| Lateral occipital | .05 | .59 | .96 | .14 | .14 | .76 | -.04 | .64 | .96 | .04 | .67 | .96 | .13 | .10 | .76 | .02 | .78 | .96 |
| Banks of the superior temporal sulcus | -.01 | .92 | .98 | .10 | .31 | .94 | -.04 | .68 | .96 | .03 | .76 | .96 | .13 | .11 | .76 | -.08 | .31 | .94 |
| *Right hemisphere* | | | | | | | | | | | | | | | | | | |
| Rostral middle frontal | .09 | .36 | .94 | .17 | .08 | .76 | -.06 | .55 | .96 | .13 | .13 | .76 | .21 | .01 | .52 | -.01 | .94 | .98 |
| Superior parietal | .04 | .65 | .96 | .03 | .78 | .96 | -.02 | .85 | .98 | .08 | .38 | .94 | .10 | .22 | .79 | .04 | .66 | .96 |
| Insula (1) | .17 | .08 | .76 | .21 | .03 | .75 | .03 | .78 | .96 | .23 | .009 | .52 | .20 | .01 | .52 | -.01 | .91 | .98 |
| Superior temporal | .08 | .39 | .94 | .13 | .17 | .76 | .02 | .85 | .98 | .11 | .20 | .76 | .12 | .13 | .76 | -.01 | .86 | .98 |
| Superior frontal (1) | .05 | .60 | .96 | .05 | .57 | .96 | -.05 | .56 | .96 | .16 | .06 | .76 | .11 | .17 | .76 | .07 | .40 | .94 |
| Precuneus (1) | .06 | .55 | .96 | .09 | .35 | .94 | .01 | .92 | .98 | .05 | .55 | .96 | .13 | .12 | .76 | -.09 | .93 | .98 |
| Precuneus (2) | -.02 | .86 | .98 | .05 | .63 | .96 | .06 | .50 | .96 | .05 | .58 | .96 | .07 | .42 | .94 | .00 | .97 | .99 |
| Insula (2) | .12 | .20 | .76 | .13 | .17 | .76 | .04 | .65 | .96 | .16 | .07 | .76 | .18 | .02 | .75 | -.04 | .62 | .96 |
| Superior frontal (2) | .03 | .74 | .96 | -.02 | .85 | .98 | .01 | .88 | .98 | .03 | .71 | .96 | .07 | .35 | .94 | .10 | .22 | .8 |
| Postcentral | -.07 | .51 | .96 | -.09 | .33 | .94 | -.02 | .83 | .98 | .00 | .99 | .99 | .00 | .99 | .99 | .00 | .99 | .99 |

Linear regression models (n=155) adjusted for age, sex, and diagnosis.

[1]For PANSS total score and negative symptom factor n=154 due to missing data in one participant.

Preprint version

### Supplementary Figure 1 – Age slopes in all significant clusters

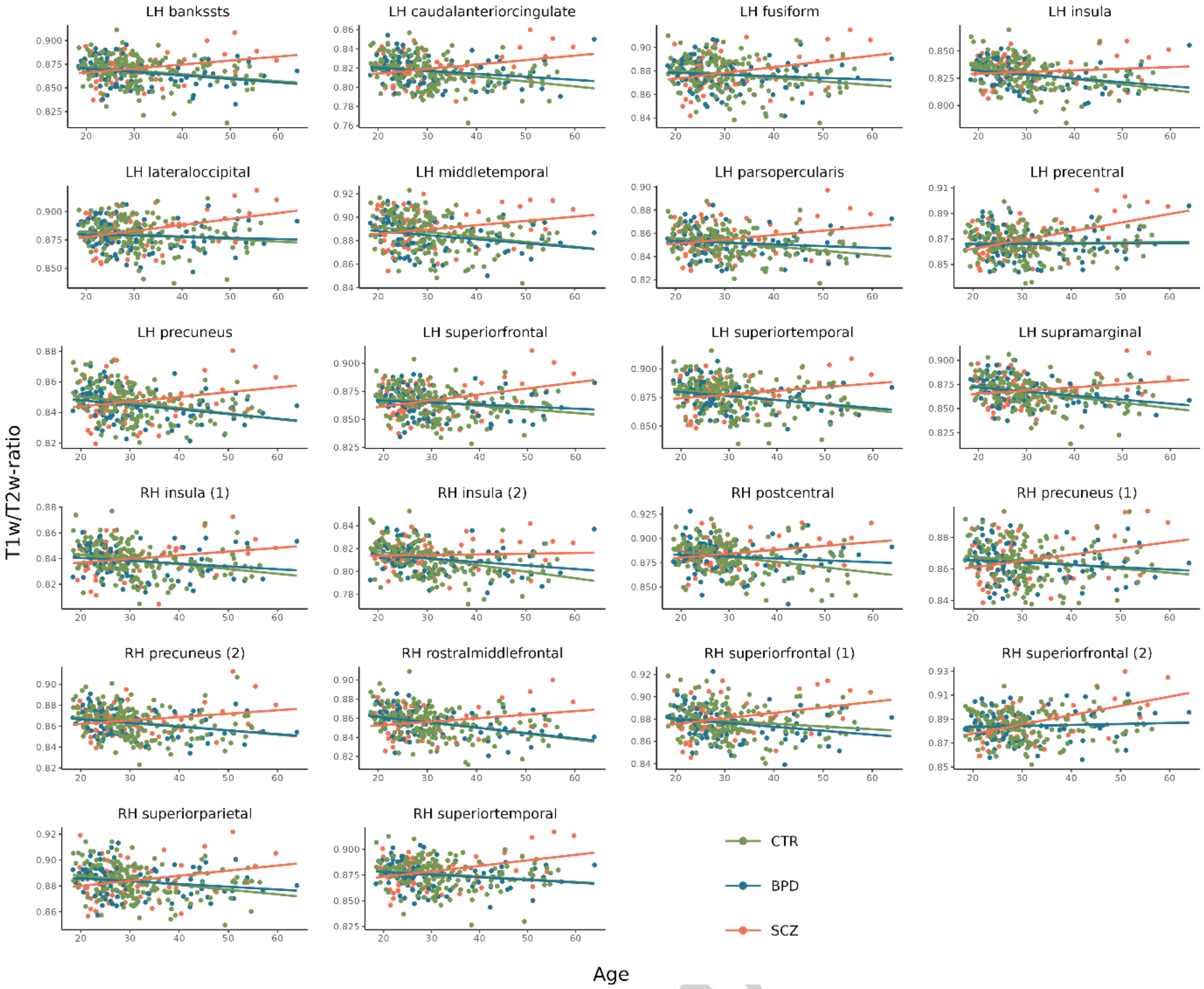


Scatterplots showing the associations with age for each significant cluster from the surface analyses for SCZ (red line), BPD (blue line) and CTR (green line). The x-axes represent age, and the y-axes represent mean T1w/T2-ratio values within the cluster.

ersion
